# Impact of COVID-19 related unemployment on increased cardiovascular disease in a high-income country: Modeling health loss, cost and equity

**DOI:** 10.1101/2020.12.15.20248284

**Authors:** Nhung Nghiem, Nick Wilson

**Affiliations:** Department of Public Health, University of Otago, Wellington 6021

**Keywords:** CVD, unemployment, COVID-19, New Zealand, health inequities, HALYs

## Abstract

**Background:** Cardiovascular disease (CVD) is a leading cause of health loss and health sector economic burdens in high-income countries. Unemployment is associated with increased risk of CVD, and so there is concern that the economic downturn associated with the COVID-19 pandemic will increase the CVD burden.

**Aims:** This modeling study aimed to quantify health loss, health cost burden and health inequities among people with CVD due to additional unemployment caused by COVID-19 pandemic-related economic disruption in one high-income country: New Zealand (NZ).

**Methods:** We adapted an established and validated multi-state life-table model for CVD in the national NZ population. We modeled indirect effects (ie, higher CVD incidence due to high unemployment rates) for various scenarios of pandemic-related unemployment projections.

**Results:** We estimated the CVD-related heath loss in NZ to range from 23,300 to 36,900 HALYs (health-adjusted life years) for the different unemployment scenarios. Health inequities for Māori (Indigenous population) were 3.7 times greater compared to non-Māori (49.9 vs 13.5 HALYs lost per 1000 people).

**Conclusions and policy implications:** Unemployment due to the COVID-19 pandemic is likely to cause significant health loss and health inequities from CVD in this high-income country. Prevention measures should be considered by governments to reduce this risk, including job creation programs and measures directed towards CVD prevention.

## 1. Introduction

The COVID-19 pandemic had infected 65 million people and caused over 1.5 million deaths globally at the time of writing (early December 2020).^(1,2)^ Unfortunately at this time, pandemic spread was still accelerating and so its final impact before widespread vaccination is used, is likely to be substantially greater.

Globally, this pandemic has disrupted international travel and domestic economies (often via the use of “lockdowns” that depress normal consumer activity and close some workplaces). As such it has depressed gross domestic product (GDP) with this figure for OECD countries being by -10.5% in the second quarter in 2020,^(3)^ and with an increase in unemployment by an absolute increment of 3.2% (8.57% in the second quarter vs 5.35% in the first quarter)^(4)^ for these countries.^(5-7)^ There is established evidence that economic recessions and depressions can increase poor health.^(8, 9)^ This includes cardiovascular disease (CVD)^(8-10)^ as per our recent review of this topic.^(11)^ In particular, one meta-analysis,^(9)^ which included 174,438 participants with a mean follow-up of 9.7 years and 1892 incident cases of CVD from 13 cohort studies, reported that increased job insecurity was associated with increased CVD incidence. Also, there is evidence that people experiencing economic hardship are at higher risk of CVD mortality.^(8, 9, 12)^ Stress and loneliness (eg, potentially exacerbated by pandemic-related lockdowns) are also known risk factors for CVD onset.^(13-15)^

According to the Global Burden of Disease Study 2017, CVD is still the leading cause of death in the world at an estimated 17.8 million deaths.^(16)^ In New Zealand (NZ), CVD makes up around 14% of all health loss nationally, and is responsible for a third of the total number of deaths annually.^(17)^ CVD is a particularly important contributor to health loss for the Indigenous Māori population^(17, 18)^ and it contributes to health inequities in NZ in terms of both ethnicity and socioeconomic position.^(19-21)^

Although NZ has had a relatively successful health sector response to the COVID-19 pandemic (with an elimination strategy^(22)^), the economic impact of the response (via lockdowns, lost revenue from international tourism and international students) has been severe.^(23, 24)^

This study therefore aimed to quantify via modeling the health loss, health cost burden and health inequities due to CVD onset as a result of unemployment caused by the COVID-19 pandemic in NZ.^(25, 26)^

## 2. Methods

We adapted an established and validated multi-state life-table model for CVD for the national NZ population.^(27-29)^ We modeled only indirect effects (ie, higher CVD incidence due to high unemployment rates) arising from COVID-19 pandemic-related unemployment. Direct effects of the COVID-19 pandemic onto CVD were not considered since the disease burden (only 25 deaths as of 15 December 2020) has been relatively low in NZ prior to the elimination of community transmission. We used various scenarios of the COVID-19 pandemic’s impact on unemployment projections in NZ.^(23, 30)^ The timeline for the pandemic impact on unemployment was for five years as per the Treasury projections, but a lifetime horizon was used for measuring benefits and costs (a five-year horizon was also implemented).

### a. Unemployment scenarios due to the COVID-19 pandemic and the response in NZ

The NZ Government stated that “the COVID-19 pandemic is a ‘once in a century’ public health shock that is also having a profound impact on economic and financial systems around the world and in NZ.”^(30)^ The Government also suggested that the impact of COVID-19 and related response measures on the NZ economy is highly uncertain. So the NZ Treasury presented four different economic scenarios based on health response measures and on key economic assumptions, including the reduction in production output in NZ, the Government’s support for households and businesses, and world real production outputs. Detailed scenarios including the baseline unemployment projection before COVID-19, the base case and alternative scenarios (ie, earlier recovery in services exports, extended border controls, and resurgence in community transmission) are presented in Table 1. The key estimates were that the peak of unemployment rates would vary from 7.7% in the most relaxed response, to 9% in the most restrictive one (Table 1). In all of the scenarios, however, the unemployment rates were still higher than that in the baseline scenario (had there never been a pandemic) which was around 4% after five years (in 2024).

**Table 1:**
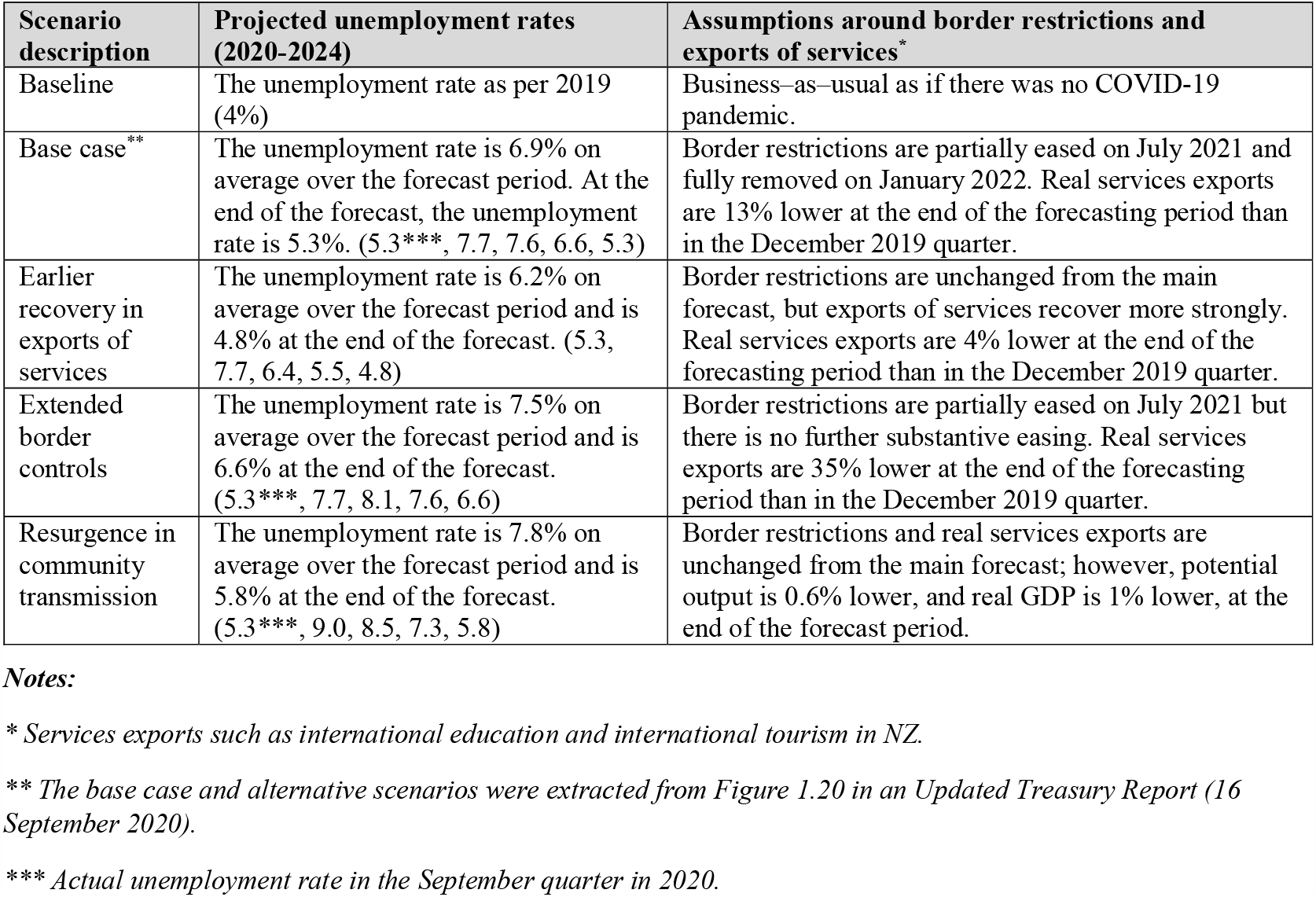
Projected unemployment scenarios in NZ as a result of the COVID-19 pandemic and response to it (extracted from a NZ Treasury Report)^(23, 30)^

### b. Calibration of the changes in unemployment rates due to the COVID-19 pandemic by age, sex and ethnicity

In the NZ Treasury Reports,^(23, 30)^ projected unemployment rates were not disaggregated by age, sex and ethnicity. We therefore calibrated the projected changes in unemployment rates annually between 2020-2024, so that it could be used directly in our model as follows.

*Data*: We averaged the unemployment rates by ethnicity (ie, Māori and non-Māori) over 2010-2014.^(31, 32)^ We also averaged the unemployment rates by sex and by age over 2010- 2014. Of note is that these rates were for people aged 15-64 years old in NZ. We used real population by age, sex and ethnicity from Census 2013 to calibrate the unemployment rates.^(33)^

#### Assumption

We assumed that the relative rate of unemployment rates by age, sex and ethnicity before the COVID-19 pandemic held. That meant that:

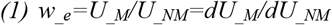

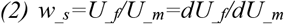

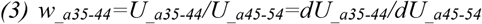

Where *U* is unemployment rate, *dU* is the absolute change in unemployment rate, *w*_*_e*_ is the weight of unemployment rate for Māori over non-Māori, *w*_*_s*_ is the weight of unemployment rate for women over men, and *w*_*_*a35-44_ is the weight of unemployment rate for people aged 35- 44 over the reference age group which is people aged 45-54 years.

#### Algorithm

From the assumptions in equations (1) to (3), we derived the absolute changes in unemployment rates by ethnicity, sex and 10-year age group, in sequence, as follows:

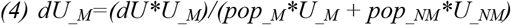

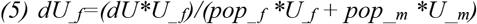

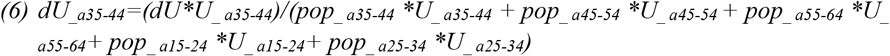

Where *pop*_*_M*_ is the real Māori population in 2013, *pop*_*_ a35-44*_ is the real population aged 35- 44 years, and so on. Of note is that if any of the assumptions in equations (1)-(3) are altered, the absolute changes derived in equations (4)-(6) are changed accordingly. Furthermore, we assumed sex and age patterns (equations (5)-(6)) followed that for the general population, not by each ethnicity. This involves a slight simplification since there are minor differences in age structure by ethnicity within the studied age-groups, eg, Māori has a younger age group. But as young people have higher unemployment rates, by using the age structure for the general population, our results were conservative towards smaller changes in absolute unemployment rates.

#### Output

The output of the calibration was absolute changes in unemployment rates (as projected by the Treasury^(23, 30)^) by ethnicity (Māori/non-Māori), sex and 10-year age group (35-64 years old) for each unemployment scenario annually (2020-2024).

### c. Associations between unemployment and, all-cause mortality and CVD incidence

In this study, we modeled health burdens from pandemic-induced unemployment in terms of CVD morbidity and mortality. We focused on only modeling the impact of unemployment on CVD incidence, and ignored other economic impacts arising from the reduction in real GDP. For the impact of unemployment on CVD incidence, we used the results from a large multi-country study by Stuckler et al in 2009.^(10)^ We considered this work to be the most robust of all the studies we identified in a recent review.^(11)^ This work reported a 0.85% relative increase in CVD incidence for a 1% relative increase in the unemployment rate in middle-aged men (or a relative risk (RR) of 1.0085 for CVD incidence (including both coronary heart disease [CHD] and stroke)). While Stuckler et al 2009 considered only CVD mortality – for modeling purposes we assumed that this impact was entirely due to increased CVD incidence and not any additional deterioration in CVD case-fatality risk (where we assumed no change in the pre-existing downward trend (Table 3)). There will be some competing mortality reasons why a 0.85% increase in incidence doesn’t perfectly align with a 0.85% increase in CVD mortality – and so we reported how the model output actually responds to the 0.85% incidence increase.

**Figure 1.**
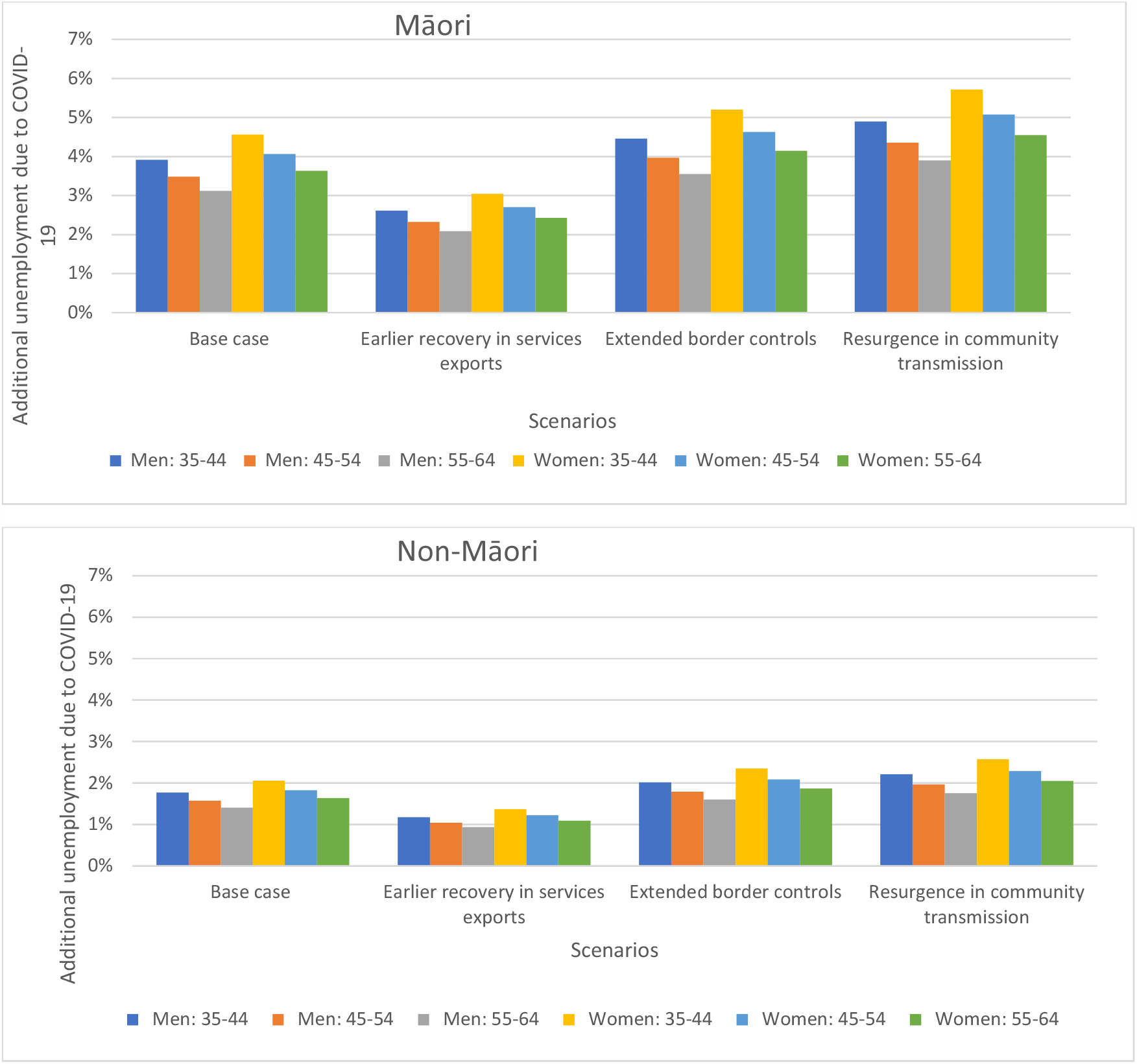
Scenarios around the COVID-19 pandemic related additional absolute unemployment rates in 2021 used in the BODE^3^ CVD model by age, sex and ethnicity in NZ^a,b,c,d^. ^*a*^ *These unemployment rates were the differences in the projected unemployment rates between having the COVID-19 pandemic (Scenarios in Table 1) and had not having the* COVID-19 *pandemic (Baseline Scenario in Table 1). They were also adjusted by the unemployment rates by age, sex and ethnicity in 2010-2014 reported by the NZ Government*. ^*b*^ *All unemployment rates followed a log-normal distribution with an assumed 20% standard deviation (due to all health responses and economic outlooks being highly uncertain)*. ^*c*^ *We only extracted unemployment rates for these age-groups as they are in typical working age age-groups, and are potentially impacted by the CVD burden. But we acknowledge this limitation further in the Discussion*. ^*d*^ *The age-groups in this Table do not exactly match the ones in Table 2, but we used the closest age group for RR and additional unemployment rate in our model*. *Note that in 2021, unemployment rates are similar across scenarios (see Table 1). Furthermore, in the Treasury economic models, the projected unemployment levels take into account the impact from GDP and other services such as impacts from the continued loss of international students and international tourists*.

We also assumed that the effect size in CVD incidence increased linearly with an increase in unemployment rate, ie, a 2% increase in <u>relative</u> unemployment rate translated to 1.7% relative increase in CVD incidence <u>rate</u> with a RR of 1.0085. The assumption of linearity is the most parsimonious assumption – especially for these relatively small changes. But it is plausible that large increases in unemployment would have normalizing effects and perhaps reduce the impact on CVD (eg, if a large increase in unemployment triggered a disproportionate government response such as a major expansion of welfare provisions).

**Table 2.** Associations between unemployment and CVD incidence used in this modeling study as per Stuckler et al 2009 (Figure 3)^(10)^

| Population group | Association of a 1% relative increase in unemployment on CVD mortality percentage changes (% and (CIs)) as per Stuckler et al 2009 | Change in annual CVD incidence (RR(CIs)) (converted to so as to use in our modeling work) |
| --- | --- | --- |
| Women, aged 35-44 | -0.14 (-1.95–1.67) | -1.0014 (-1.0195–1.0167) |
| 45-59 | 0.10 (-1.09–1.30) | 1.0010 (-1.0109–1.0130) |
| 60-64 | 0.23 (-0.22–0.68) | 1.0023 (-1.0022–1.0068) |
| Men, aged 35-44 | 0.85 (0.06–1.64) | 1.0085 (1.0006–1.0164) |
| 45-59 | 0.48 (-0.06–1.02) | 1.0048 (-1.0006–1.0102) |
| 60-64 | 0.38 (-0.16–0.91) | 1.0038 (-1.0016–1.0091) |

**Table 3.** Key epidemiology and economic parameters for the established BODE^3^ CVD model^(27-29)^

| Parameters | Value | Heterogeneity | Uncertainty |
| --- | --- | --- | --- |
| All-cause mortality and time trends | As per the BODE<sup>3</sup> CVD model.<sup>(27-29)</sup> Trends for all-cause mortality were made consistent with long-run mortality trends for NZ (annual 2.25% mortality decline for Māori and 1.75% per annum for non-Māori). Trends were modeled out to 2026 (ie, for 15 years), with no subsequent decline for both ethnic groupings thereafter. | By ethnicity (Māori/non-Māori), sex and age | No |
| Baseline CVD incidence/morbidity /mortality and time trends | As per the BODE<sup>3</sup> CVD model.<sup>(27-29)</sup> As per the NZ Burden of Disease Study (NZBDS)<sup>(34)</sup> we assumed a continued decline in incidence rates for both CHD and stroke of 2.0% annually, and also a 2.0% annual reduction in case fatality so that it reflects improved treatment and management. | By ethnicity (Māori/non-Māori), sex and age | Uncertainty: ±5% SD, log-normal distribution. |
| Background health system costs for all citizens (adjusted for CHD and stroke costs) | As per BODE<sup>3</sup> costing methods<sup>(35)</sup> | By sex and age | Uncertainty: ±10% SD, log-normal distribution. |
| Unemployment duration | 5 years as per the NZ Treasury report<sup>(23)</sup> | No | No, but we conducted a sensitivity analysis whereby unemployment persists for 10 years. |
| Unemployment rates | As per the NZ Treasury's report over 2020-2024 (Table 1) | By ethnicity (Māori/non-Māori), sex and age | All unemployment rates followed a log-normal distribution with an assumed 20% standard deviation (due to all health responses and economic outlooks being highly uncertain). |
| Discount rate | 3% for both health losses and costs as per standard BODE<sup>3</sup> methods | No | No |
| Perspective | Health system as per standard BODE<sup>3</sup> methods | No | No |
| Time horizon | Lifetime (until died or 110 years old) | No | No |

### d. Key model parameters: epidemiology, economic and unemployment

We presented all epidemiology and economic key parameters as per the established and validated multi-state life-table BODE^3^ model for CVD for the national NZ population in Table 3.^(27-29)^

We calculated unemployment rates by age, sex and ethnicity used in the BODE^3^ CVD model for each economic scenario listed in Table 1. These were the differences in the projected unemployment rates between the pre-COVID-19 baseline and after the appearance of the COVID-19 pandemic (Baseline Scenario in Table 1). All unemployment rates followed a log- normal distribution with an assumed 20% standard deviation (due to all health responses and economic outlooks being highly uncertain).

Of note is that due to the design of the BODE^3^ CVD model, we modeled the unemployment impacts due to the COVID-19 pandemic as if they were happening in the 2011-2015 period.

### e. Additional scenarios and sensitivity analysis

Further to four economic scenarios as mentioned above, we also modeled the following sensitivity analyses:

A. Māori CVD epidemiology parameters as per non-Māori to avoid penalizing Māori due to lower life expectancy and higher comorbidity,
B. Unemployment persists for longer (ie, 10 years),
C. No reduction in CVD trend over time (eg, on the assumption that the long-term downward trend in CVD is halted by the obesity epidemic – as seen in some high-income countries).^(36)^
D. Zero percent discount rate, and
E. Six percent discount rate.

## 3. Results

Table 4 shows the estimated CVD-related health loss (in HALYs) for the base case (most likely scenario) (3% discount rate for the remaining life of the NZ population alive in 2011) for various COVID-19 pandemic induced unemployment scenarios (base case, early recovery and extended border control). The health loss for the base-case was estimated at -30,300 HALYs (uncertainty interval: -66,700 to 4,100). However, only 10% of the loss (−3,010 HALYs) was accumulated in the first five years and more than 50% (−17,770 HALYs) in the 20 years in the future. The estimated CVD-related heath loss in NZ ranged from 23,300 (early recovery in export and services) to 36,900 lost HALYs (resurgence in community transmission) for the different unemployment scenarios. Of which, health loss for Māori ranged between -7,700 HALYs and -13,000, and for non-Māori from -15,700 to -26,600.

**Table 4.**
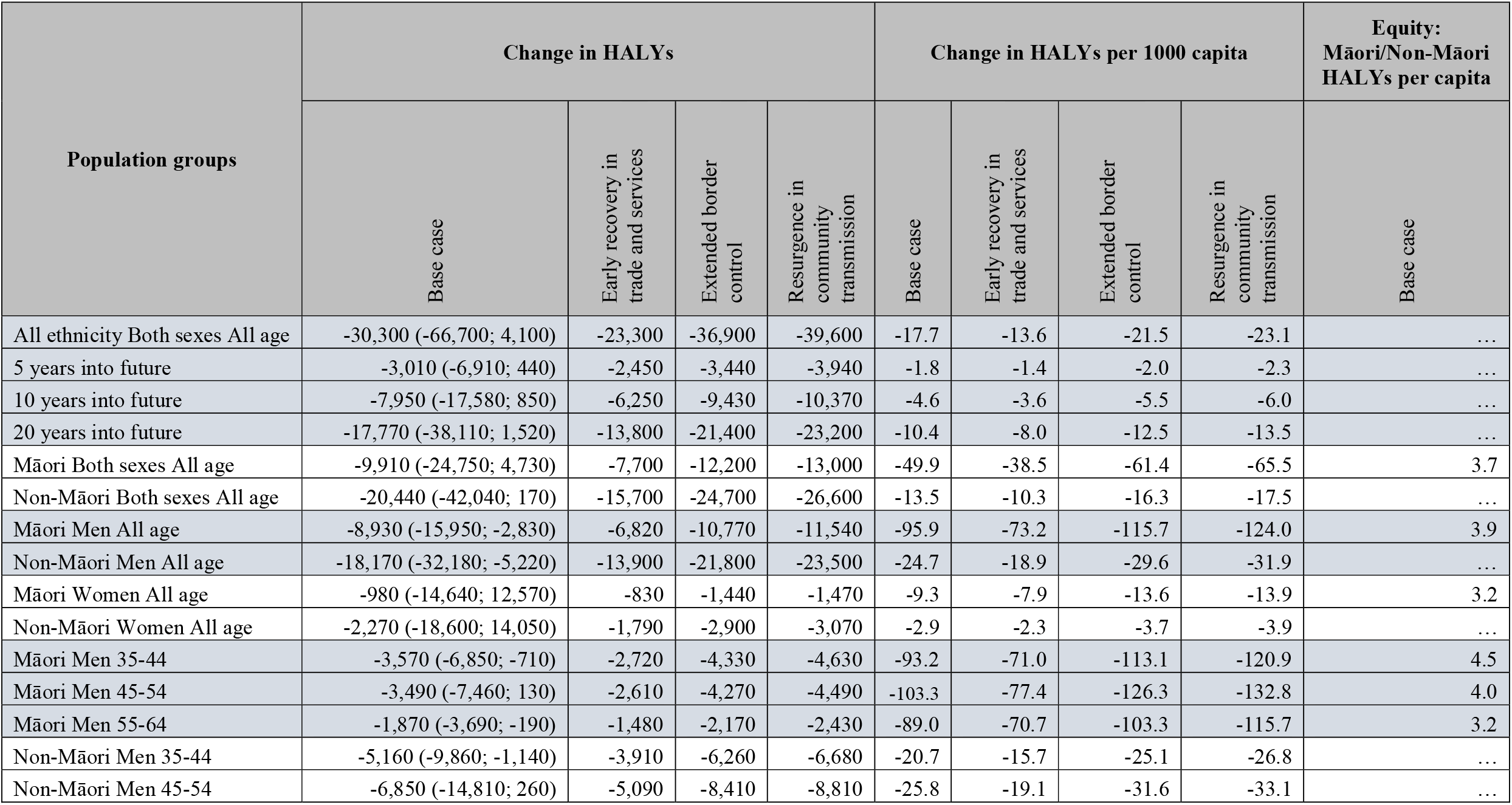
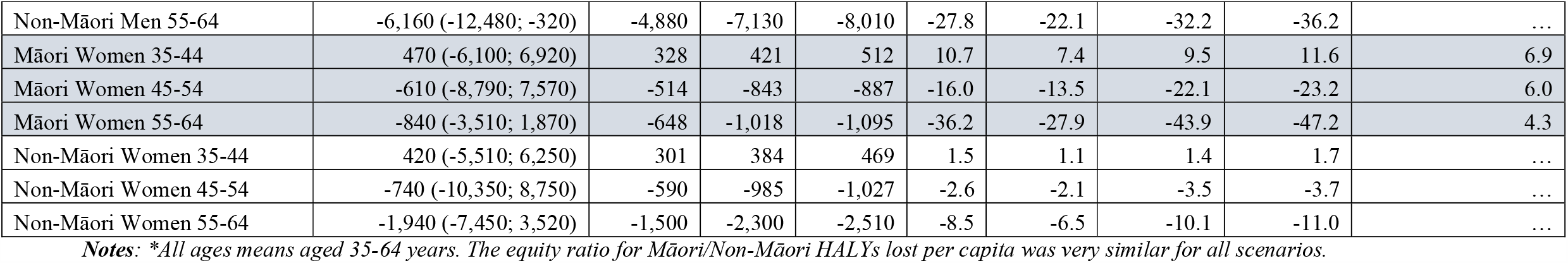
Health loss (in HALYs) results for the base case (most likely unemployment scenario) (3% discount rate for the remaining life of the NZ population alive in 2011) for various COVID-19 pandemic-induced unemployment scenarios from the NZ Treasury.

| Population groups | Change in HALYs |  |  |  | Change in HALYs per 1000 capita |  |  |  | Equity:<br>Māori/Non-Māori<br>HALYs per capita |
| --- | --- | --- | --- | --- | --- | --- | --- | --- | --- |
|  | Base case | Early recovery in<br>trade and services | Extended border<br>control | Resurgence in<br>community<br>transmission | Base case | Early recovery in<br>trade and services | Extended border<br>control | Resurgence in<br>community<br>transmission | Base case |
| All ethnicity Both sexes All age | -30,300 (-66,700; 4,100) | -23,300 | -36,900 | -39,600 | -17.7 | -13.6 | -21.5 | -23.1 | ... |
| 5 years into future | -3,010 (-6,910; 440) | -2,450 | -3,440 | -3,940 | -1.8 | -1.4 | -2.0 | -2.3 | ... |
| 10 years into future | -7,950 (-17,580; 850) | -6,250 | -9,430 | -10,370 | -4.6 | -3.6 | -5.5 | -6.0 | ... |
| 20 years into future | -17,770 (-38,110; 1,520) | -13,800 | -21,400 | -23,200 | -10.4 | -8.0 | -12.5 | -13.5 | ... |
| Māori Both sexes All age | -9,910 (-24,750; 4,730) | -7,700 | -12,200 | -13,000 | -49.9 | -38.5 | -61.4 | -65.5 | 3.7 |
| Non-Māori Both sexes All age | -20,440 (-42,040; 170) | -15,700 | -24,700 | -26,600 | -13.5 | -10.3 | -16.3 | -17.5 | ... |
| Māori Men All age | -8,930 (-15,950; -2,830) | -6,820 | -10,770 | -11,540 | -95.9 | -73.2 | -115.7 | -124.0 | 3.9 |
| Non-Māori Men All age | -18,170 (-32,180; -5,220) | -13,900 | -21,800 | -23,500 | -24.7 | -18.9 | -29.6 | -31.9 | ... |
| Māori Women All age | -980 (-14,640; 12,570) | -830 | -1,440 | -1,470 | -9.3 | -7.9 | -13.6 | -13.9 | 3.2 |
| Non-Māori Women All age | -2,270 (-18,600; 14,050) | -1,790 | -2,900 | -3,070 | -2.9 | -2.3 | -3.7 | -3.9 | ... |
| Māori Men 35-44 | -3,570 (-6,850; -710) | -2,720 | -4,330 | -4,630 | -93.2 | -71.0 | -113.1 | -120.9 | 4.5 |
| Māori Men 45-54 | -3,490 (-7,460; 130) | -2,610 | -4,270 | -4,490 | -103.3 | -77.4 | -126.3 | -132.8 | 4.0 |
| Māori Men 55-64 | -1,870 (-3,690; -190) | -1,480 | -2,170 | -2,430 | -89.0 | -70.7 | -103.3 | -115.7 | 3.2 |
| Non-Māori Men 35-44 | -5,160 (-9,860; -1,140) | -3,910 | -6,260 | -6,680 | -20.7 | -15.7 | -25.1 | -26.8 | ... |
| Non-Māori Men 45-54 | -6,850 (-14,810; 260) | -5,090 | -8,410 | -8,810 | -25.8 | -19.1 | -31.6 | -33.1 | ... |
|  | Base case | Early recovery in<br>trade and services | Extended border<br>control | Resurgence in<br>community<br>transmission | Base case | Early recovery in<br>trade and services | Extended border<br>control | Resurgence in<br>community<br>transmission | Base case |
| Non-Māori Men 55-64 | -6,160 (-12,480; -320) | -4,880 | -7,130 | -8,010 | -27.8 | -22.1 | -32.2 | -36.2 | ... |
| Māori Women 35-44 | 470 (-6,100; 6,920) | 328 | 421 | 512 | 10.7 | 7.4 | 9.5 | 11.6 | 6.9 |
| Māori Women 45-54 | -610 (-8,790; 7,570) | -514 | -843 | -887 | -16.0 | -13.5 | -22.1 | -23.2 | 6.0 |
| Māori Women 55-64 | -840 (-3,510; 1,870) | -648 | -1,018 | -1,095 | -36.2 | -27.9 | -43.9 | -47.2 | 4.3 |
| Non-Māori Women 35-44 | 420 (-5,510; 6,250) | 301 | 384 | 469 | 1.5 | 1.1 | 1.4 | 1.7 | ... |
| Non-Māori Women 45-54 | -740 (-10,350; 8,750) | -590 | -985 | -1,027 | -2.6 | -2.1 | -3.5 | -3.7 | ... |
| Non-Māori Women 55-64 | -1,940 (-7,450; 3,520) | -1,500 | -2,300 | -2,510 | -8.5 | -6.5 | -10.1 | -11.0 | ... |

*Notes: \*All ages means aged 35-64 years. The equity ratio for Māori/Non-Māori HALYs lost per capita was very similar for all scenarios.*

For the base case (best-estimate) scenario, health inequities for Māori (Indigenous population) were 3.7 times greater than for non-Māori (49.9 vs 13.5 HALYs lost per 1000 people). Māori men suffered the most health loss per capita (95.9 HALYs per 1000 people); however, the worst inequity impacts between Māori and non-Māori were seen in women aged 45-54 years across all scenarios (six fold difference). Inequities were exacerbated much more than the sum of existing inequities in CVD burden and unemployment, ie, in some groups, the CVD burden for Māori was increased to six times higher than that of non-Māori (compared to a double unemployment rate or CVD burden). The overall pattern of inequity by ethnicity, age and sex was similar among various economic scenarios.

Table 5 presents the additional health costs incurred to the NZ health system due to the higher CVD burden. The estimates ranged between NZ$276 million (m) to 458m in 2011 value (NZ$303m to 503m or US$209m to 346m in 2019 values) for the various unemployment scenarios. Similar to the health burden, 10% of the additional health cost was accumulated in the first five years and more than 60% in the 20 years in the future.

**Table 5.** Additional health system costs for the base case (most likely unemployment scenario) (3% discount rate for the remaining life of the NZ population alive in 2011)

| Groups | Additional health system costs (NZ\$ million [m]) | | | | Additional health system costs (NZ\$ m) per 1000 capita | | | |
| --- | --- | --- | --- | --- | --- | --- | --- | --- |
|  | Base case | Early recovery in trade and services | Extended border control | Resurgence in community transmission | Base case | Early recovery in trade and services | Extended border control | Resurgence in community transmission |
| All ethnicity Both sexes All age | 354 (-78; 824) | 276 | 429 | 458.0 | 0.21 | 0.16 | 0.25 | 0.27 |
| 5 years into future | 234 (-6; 501) | 187 | 276 | 301.0 | 0.14 | 0.11 | 0.16 | 0.18 |
| 10 years into future | 324 (-26; 703) | 254 | 388 | 418.0 | 0.19 | 0.15 | 0.23 | 0.24 |
| 20 years into future | 402 (-63; 902) | 312 | 487 | 520.0 | 0.23 | 0.18 | 0.28 | 0.30 |

The CVD-related health loss, inequities and health costs were similar in term of relative patterns across scenario analyses and sensitivity analyses (Supplementary Tables 1–5). The equity scenario (Scenario #A), where Māori CVD epidemiology parameters were assumed to be as per non-Māori, suggested more health loss (up to -13,200 HALYs) and more inequity for Māori population (up to a ratio of Māori/non-Māori HALYs of 7.1). Unemployment persisting for 10 years (#B) suggested the most health loss (−57,100 HALYs), and 6% discount rates (#E) resulted in the smallest health loss (−18,500 HALYs).

## 4. Discussion and conclusions

### a. Main results and interpretation

In this modeling study, we estimated the CVD-related heath loss in NZ to range from 23,300 to 36,900 HALYs for the different unemployment scenarios. For the base case (best-estimate) scenario, health inequities for Māori were 3.7 times greater. While Māori men suffered the most health loss per capita, the worst inequity impacts between Māori and non-Māori were seen in women aged 45-54 years across all scenarios (six fold difference). Inequities were exacerbated much more than the sum of existing inequities in CVD burden and unemployment, ie, in some groups, Māori’s CVD burden was increased to six times higher than that of non-Māori (compared to a double unemployment rate or CVD burden). The additional health costs due to higher CVD burden were also substantial (at NZ$276m to 458m in 2011 values) for the various unemployment scenarios.

### b. Study strengths

Our study has several strengths as follows. First, we used a validated disease model that has been previously applied in the NZ context. Second, we followed economic scenarios projected by the NZ Treasury with valid assumptions on border controls, international tourism and education for NZ. Third, as the COVID-19 pandemic response in NZ was among the strictest^(37)^ and one of the most successful as of December 2020,^(38)^ our analysis reflects a pandemic related impact that is relatively independent of the direct disease impacts (ie, it reflects unemployment from the indirect impacts of the NZ response and international impacts impacting on trade). Finally, our study benefited from detailed unemployment data by age, sex and ethnicity in NZ.

### c. Study limitations

We did not consider that all-cause mortality and CVD mortality can also change due to behavioral changes vs supply changes (tobacco, fast food, alcohol, and physical activity level) as a result of the COVID-19 pandemic restrictions. We assumed that the background trend in case fatality risk before the COVID-19 pandemic still held. While it is possible that there is an effect of recessions on poorer health care delivery care (that might increase case fatality risk in those with CVD), in the absence of data we assumed no such additional impact. Also if a recession lowers the affordability of tobacco and alcohol – then there might be non-linear social contagion impacts as whole cohorts of people reduce consumption of these products in social settings (eg, going out less to bars and restaurants). But we had inadequate data on such impacts to consider these in this research. If increased unemployment-induced poverty resulted in less affordable tobacco and less expenditure on junk food (generally more expensive than home-cooked food in NZ^(39)^),– then those factors would tend to reduce CVD. Finally, we ignored people aged 64+ who are in retirement age (although around 20% of this group are still in formal employment in NZ).

### d. Potential policy implications

This modeling work suggests that unemployment due to the COVID-19 pandemic is likely to cause significant health loss and health costs from CVD in NZ. Furthermore, this burden exacerbates the health inequities in CVD for Māori due to higher background risks of CVD, and higher unemployment rates. Prevention measures should be considered by central and local governments to reduce this risk, including job creation programs and measures directed towards CVD prevention eg, enhanced progress towards a smokefree country and reducing the hazardous processed food environment as we discuss elsewhere.^(11)^

### e. Further research work

Further work could be done on modeling health inequity, health burden and health cost impacts of COVID-19-related unemployment by occupation. It may also be worth modeling unemployment interventions. Examples could be assuming that a government could implement a job creation package, such as via promoting tourism and education to international visitors and students, so that an absolute 1% of additional jobs could be created over the period 2020-2024. In addition, a modeled intervention could assume that the NZ Government could target job creation for Māori so that the absolute increases in unemployment rates of Māori were reduced to those of non-Māori. Some of these new jobs could have substantial co-benefits eg, additional tree planting on farmed hill country has benefits in terms of carbon sequestration, enhanced biodiversity, erosion prevention, flood prevention and improved water quality.

To conclude, this modeling work suggests that unemployment due to the COVID-19 pandemic is likely to cause significant health loss, health costs and exacerbate inequities from CVD in this high-income country. Prevention measures should be considered by governments to reduce this risk, including job creation programs and measures directed towards CVD prevention (eg, enhanced progress towards a smokefree country and reducing the hazardous processed food environment).

## Data Availability

Sharing of anonymized cohort data with other researchers or official agencies of the other epidemiological and costing data will generally be possible on request from the authors (pending approval of the relevant official agencies).

## Acknowledgement

NN and NW were supported by the Health Research Council of NZ (grant 10/248 and grant 16/443). The authors thank Dr Anja Mizdrak for helpful comments on the design of the study and modeling parameters.

## Author contribution

The study was designed by NN and NW. NN, NW led the data collection and parameter specification. NN adapted the CVD version of the model and ran the analyses. NN drafted the manuscript. NW substantially edited and revised the manuscript. All authors approved the final manuscript.

### Ethics Approval

Approval for use of anonymized administrative data as part of the BODE^3^ Programme has been granted by the Health and Disability Ethics Committees (reference number H13/049).

---

**Supplementary Table 1.** Health loss (in HALYs) results for the Equity scenario (Scenario analysis #A) (3% discount rate for the remaining life of the NZ population alive in 2011) for various unemployment scenarios.

| Groups | Change in HALYs |  |  |  | Change in HALYs per 1000 capita |  |  |  | Equity:<br>Māori/Non-Māori HALYs<br>per capita |
| --- | --- | --- | --- | --- | --- | --- | --- | --- | --- |
|  | Base case | Early recovery in<br>trade and services | Extended border<br>control | Resurgence in<br>community<br>transmission | Base case | Early recovery in<br>trade and services | Extended border<br>control | Resurgence in<br>community<br>transmission | Base case |
| All ethnicity Both sexes All age | -31,200 | -24,000 | -37,900 | -40,600 | -18.2 | -14.0 | -22.1 | -23.7 | ... |
| 5 years into future | -3,030 | -2,460 | -3,450 | -3,960 | -1.8 | -1.4 | -2.0 | -2.3 | ... |
| 10 years into future | -8,050 | -6,320 | -9,540 | -10,490 | -4.7 | -3.7 | -5.6 | -6.1 | ... |
| 20 years into future | -18,100 | -14,000 | -21,800 | -23,600 | -10.6 | -8.2 | -12.7 | -13.8 | ... |
| Māori Both sexes All age | -10,800 | -8,300 | -13,200 | -14,100 | -54.4 | -41.8 | -66.5 | -71.0 | 4.0 |
| Non-Māori Both sexes All age | -20,400 | -15,700 | -24,700 | -26,600 | -13.5 | -10.4 | -16.3 | -17.5 | ... |
| Māori Men All age | -9,630 | -7,390 | -11,670 | -12,510 | -103.4 | -79.4 | -125.3 | -134.4 | 4.2 |
| Non-Māori Men All age | -18,000 | -13,900 | -21,800 | -23,500 | -24.4 | -18.9 | -29.6 | -31.9 | ... |
| Māori Women All age | -1,210 | -900 | -1,540 | -1,580 | -11.5 | -8.5 | -14.6 | -15.0 | 3.8 |
| Non-Māori Women All age | -2,350 | -1,790 | -2,900 | -3,070 | -3.0 | -2.3 | -3.7 | -3.9 | ... |
| Māori Men 35-44 | -3,820 | -2,920 | -4,650 | -4,970 | -99.7 | -76.2 | -121.4 | -129.8 | 4.8 |
| Māori Men 45-54 | -3,760 | -2,840 | -4,640 | -4,880 | -111.2 | -84.0 | -137.3 | -144.4 | 4.4 |
| Māori Men 55-64 | -2,050 | -1,630 | -2,380 | -2,660 | -97.6 | -77.6 | -113.3 | -126.7 | 3.5 |
| Non-Māori Men 35-44 | -5,130 | -3,910 | -6,260 | -6,680 | -20.6 | -15.7 | -25.1 | -26.8 | ... |
| Non-Māori Men 45-54 | -6,760 | -5,090 | -8,410 | -8,810 | -25.4 | -19.1 | -31.6 | -33.1 | ... |
| Non-Māori Men 55-64 | -6,150 | -4,880 | -7,130 | -8,010 | -27.8 | -22.0 | -32.2 | -36.2 | ... |
| Māori Women 35-44 | 417 | 348 | 447 | 543 | 9.5 | 7.9 | 10.1 | 12.3 | 7.1 |
| Māori Women 45-54 | -724 | -547 | -897 | -944 | -19.0 | -14.3 | -23.5 | -24.7 | 6.7 |
|  | Base case | Early recovery in<br>trade and services | Extended border<br>control | Resurgence in<br>community<br>transmission | Base case | Early recovery in<br>trade and services | Extended border<br>control | Resurgence in<br>community<br>transmission | Base case |
| Māori Women 55-64 | -904 | -696 | -1,094 | -1,177 | -39.0 | -30.0 | -47.2 | -50.7 | 4.6 |
| Non-Māori Women 35-44 | 361 | 301 | 384 | 469 | 1.3 | 1.1 | 1.4 | 1.7 | ... |
| Non-Māori Women 45-54 | -788 | -590 | -985 | -1,027 | -2.8 | -2.1 | -3.5 | -3.7 | ... |
| Non-Māori Women 55-64 | -1,930 | -1,500 | -2,300 | -2,510 | -8.4 | -6.6 | -10.1 | -11.0 | ... |

*Notes: All analyses in this table were implemented without uncertainty. Furthermore, please refer to the Additional scenarios and sensitivity analyses in the Methods section for changes in assumptions compared to the base-case analysis. The equity ratio for Māori/Non-Māori HALYs lost per capita was very similar for all scenarios.*

**Supplementary Table 2.** Health loss (in HALYs) results for the base case (unemployment persists for 10 years, Scenario analysis #B) for various unemployment scenarios.

| Groups | Change in HALYs |  |  |  | Change in HALYs per 1000 capita |  |  |  | Equity:<br>Māori/Non-Māori HALYs<br>per capita |
| --- | --- | --- | --- | --- | --- | --- | --- | --- | --- |
|  | Base case | Early recovery in<br>trade and services | Extended border<br>control | Resurgence in<br>community<br>transmission | Base case | Early recovery in<br>trade and services | Extended border<br>control | Resurgence in<br>community<br>transmission | Base case |
| All ethnicity Both sexes All age | -57,100 | -44,300 | -69,100 | -73,300 | -33.3 | -25.8 | -40.3 | -42.7 | ... |
| 5 years into future | -3,100 | -2,510 | -3,530 | -4,040 | -1.8 | -1.5 | -2.1 | -2.4 | ... |
| 10 years into future | -10,800 | -8,470 | -12,890 | -13,970 | -6.3 | -4.9 | -7.5 | -8.1 | ... |
| 20 years into future | -29,600 | -23,000 | -35,800 | -38,100 | -17.3 | -13.4 | -20.9 | -22.2 | ... |
| Māori Both sexes All age | -18,700 | -14,500 | -22,700 | -24,000 | -94.2 | -73.0 | -114.3 | -120.8 | 3.7 |
| Non-Māori Both sexes All age | -38,300 | -29,800 | -46,400 | -49,300 | -25.3 | -19.7 | -30.6 | -32.5 | ... |
| Māori Men All age | -17,260 | -13,470 | -20,760 | -22,090 | -185.4 | -144.7 | -223.0 | -237.3 | 3.9 |
| Non-Māori Men All age | -34,900 | -27,200 | -42,000 | -44,800 | -47.4 | -36.9 | -57.0 | -60.8 | ... |
| Māori Women All age | -1,480 | -1,050 | -1,980 | -1,920 | -14.0 | -10.0 | -18.8 | -18.2 | 3.2 |
| Non-Māori Women All age | -3,430 | -2,560 | -4,350 | -4,450 | -4.4 | -3.3 | -5.6 | -5.7 | ... |
| Māori Men 35-44 | -7,980 | -6,260 | -9,550 | -10,200 | -208.4 | -163.4 | -249.3 | -266.3 | 4.3 |
| Māori Men 45-54 | -6,720 | -5,180 | -8,210 | -8,600 | -198.8 | -153.3 | -242.9 | -254.4 | 3.7 |
| Māori Men 55-64 | -2,560 | -2,030 | -2,990 | -3,290 | -121.9 | -96.7 | -142.4 | -156.7 | 3.2 |
| Non-Māori Men 35-44 | -12,050 | -9,430 | -14,450 | -15,440 | -48.4 | -37.9 | -58.0 | -62.0 | ... |
| Non-Māori Men 45-54 | -14,350 | -11,020 | -17,630 | -18,430 | -54.0 | -41.4 | -66.3 | -69.3 | ... |
| Non-Māori Men 55-64 | -8,500 | -6,740 | -9,940 | -10,980 | -38.4 | -30.4 | -44.9 | -49.6 | ... |
| Māori Women 35-44 | 999 | 844 | 1,073 | 1,274 | 22.7 | 19.1 | 24.3 | 28.9 | 7.2 |
|  | Base case | Early recovery in<br>trade and services | Extended border<br>control | Resurgence in<br>community<br>transmission | Base case | Early recovery in<br>trade and services | Extended border<br>control | Resurgence in<br>community<br>transmission | Base case |
| Māori Women 45-54 | -1,378 | -1,058 | -1,690 | -1,771 | -36.1 | -27.7 | -44.2 | -46.4 | 5.6 |
| Māori Women 55-64 | -1,098 | -834 | -1,358 | -1,424 | -47.3 | -35.9 | -58.5 | -61.4 | 4.3 |
| Non-Māori Women 35-44 | 853 | 726 | 905 | 1,088 | 3.1 | 2.7 | 3.3 | 4.0 | ... |
| Non-Māori Women 45-54 | -1,798 | -1,379 | -2,214 | -2,311 | -6.4 | -4.9 | -7.9 | -8.3 | ... |
| Non-Māori Women 55-64 | -2,490 | -1,900 | -3,040 | -3,230 | -10.9 | -8.3 | -13.3 | -14.1 | ... |

*Notes: All analyses in this table were implemented without uncertainty. Furthermore, please refer to the Additional scenarios and sensitivity analyses in the Methods section for changes in assumptions compared to the base-case analysis. The equity ratio for Māori/Non-Māori HALYs lost per capita was very similar for all scenarios.*

**Supplementary Table 3.** Health loss (in HALYs) results for the base case (CVD incidence trend level-off, Scenario analysis #C) for various unemployment scenarios.

| Groups | Change in HALYs |  |  |  | Change in HALYs per 1000 capita |  |  |  | Equity: Māori/Non-Māori HALYs per capita |
| --- | --- | --- | --- | --- | --- | --- | --- | --- | --- |
|  | Base case | Early recovery in trade and services | Extended border control | Resurgence in community transmission | Base case | Early recovery in trade and services | Extended border control | Resurgence in community transmission | Base case |
| All ethnicity Both sexes All age | -31,200 | -23,800 | -38,000 | -40,600 | -18.2 | -13.9 | -22.2 | -23.7 | ... |
| 5 years into future | -3,120 | -2,530 | -3,570 | -4,070 | -1.8 | -1.5 | -2.1 | -2.4 | ... |
| 10 years into future | -8,250 | -6,460 | -9,820 | -10,750 | -4.8 | -3.8 | -5.7 | -6.3 | ... |
| 20 years into future | -18,400 | -14,200 | -22,200 | -23,900 | -10.7 | -8.3 | -12.9 | -13.9 | ... |
| Māori Both sexes All age | -10,300 | -7,800 | -12,600 | -13,400 | -51.9 | -39.3 | -63.4 | -67.5 | 3.8 |
| Non-Māori Both sexes All age | -20,900 | -16,000 | -25,400 | -27,200 | -13.8 | -10.6 | -16.7 | -17.9 | ... |
| Māori Men All age | -9,130 | -6,990 | -11,120 | -11,870 | -98.1 | -75.1 | -119.4 | -127.5 | 3.9 |
| Non-Māori Men All age | -18,500 | -14,200 | -22,400 | -24,000 | -25.1 | -19.3 | -30.4 | -32.6 | ... |
| Māori Women All age | -1,160 | -860 | -1,490 | -1,520 | -11.0 | -8.2 | -14.1 | -14.4 | 3.5 |
| Non-Māori Women All age | -2,420 | -1,830 | -2,990 | -3,150 | -3.1 | -2.3 | -3.8 | -4.0 | ... |
| Māori Men 35-44 | -3,670 | -2,800 | -4,490 | -4,780 | -95.8 | -73.1 | -117.2 | -124.8 | 4.5 |
| Māori Men 45-54 | -3,550 | -2,680 | -4,410 | -4,620 | -105.0 | -79.3 | -130.5 | -136.7 | 4.0 |
| Māori Men 55-64 | -1,910 | -1,510 | -2,220 | -2,480 | -91.0 | -71.9 | -105.7 | -118.1 | 3.2 |
| Non-Māori Men 35-44 | -5,280 | -4,010 | -6,470 | -6,870 | -21.2 | -16.1 | -26.0 | -27.6 | ... |
| Non-Māori Men 45-54 | -6,930 | -5,200 | -8,670 | -9,030 | -26.1 | -19.6 | -32.6 | -34.0 | ... |
| Non-Māori Men 55-64 | -6,260 | -4,950 | -7,270 | -8,140 | -28.3 | -22.4 | -32.8 | -36.8 | ... |
| Māori Women 35-44 | 404 | 336 | 433 | 525 | 9.2 | 7.6 | 9.8 | 11.9 | 6.7 |

| Groups | Change in HALYs |  |  |  | Change in HALYs per 1000 capita |  |  |  | Equity:<br>Māori/Non-Māori HALYs<br>per capita |
| --- | --- | --- | --- | --- | --- | --- | --- | --- | --- |
|  | Base case | Early recovery<br>in trade and<br>services | Extended border<br>control | Resurgence in<br>community<br>transmission | Base case | Early recovery<br>in trade and<br>services | Extended border<br>control | Resurgence in<br>community<br>transmission | Base case |
| Māori Women 45-54 | -702 | -529 | -874 | -915 | -18.4 | -13.8 | -22.9 | -24.0 | 6.3 |
| Māori Women 55-64 | -865 | -664 | -1,051 | -1,127 | -37.3 | -28.6 | -45.3 | -48.6 | 4.3 |
| Non-Māori Women 35-44 | 369 | 308 | 394 | 480 | 1.4 | 1.1 | 1.4 | 1.8 | ... |
| Non-Māori Women 45-54 | -812 | -606 | -1,020 | -1,058 | -2.9 | -2.2 | -3.6 | -3.8 | ... |
| Non-Māori Women 55-64 | -1,970 | -1,530 | -2,360 | -2,570 | -8.6 | -6.7 | -10.3 | -11.2 | ... |

*Notes: All analyses in this table were implemented without uncertainty. Furthermore, please refer to the Additional scenarios and sensitivity analyses in the Methods section for changes in assumptions compared to the base-case analysis. The equity ratio for Māori/Non-Māori HALYs lost per capita was very similar for all scenarios.*

**Supplementary Table 4.** Health loss (in HALYs) results for the base case (0% discount rate, Sensitivity analysis #D) for various unemployment scenarios.

| Groups | Change in HALYs |  |  |  | Change in HALYs per 1000 capita |  |  |  | Equity:<br>Māori/Non-Māori HALYs<br>per capita |
| --- | --- | --- | --- | --- | --- | --- | --- | --- | --- |
|  | Base case | Early recovery<br>in trade and<br>services | Extended border<br>control | Resurgence in<br>community<br>transmission | Base case | Early recovery<br>in trade and<br>services | Extended border<br>control | Resurgence in<br>community<br>transmission | Base case |
| All ethnicity Both sexes All age | -56,200 | -42,800 | -68,700 | -73,100 | -32.8 | -25.0 | -40.1 | -42.6 | ... |
| 5 years into future | -3,270 | -2,650 | -3,730 | -4,260 | -1.9 | -1.5 | -2.2 | -2.5 | ... |
| 10 years into future | -9,380 | -7,340 | -11,140 | -12,220 | -5.5 | -4.3 | -6.5 | -7.1 | ... |
| 20 years into future | -24,600 | -19,000 | -29,700 | -32,000 | -14.3 | -11.1 | -17.3 | -18.7 | ... |
| Māori Both sexes All age | -17,900 | -13,600 | -22,000 | -23,300 | -90.1 | -68.5 | -110.8 | -117.3 | 3.6 |
| Non-Māori Both sexes All age | -38,300 | -29,200 | -46,700 | -49,800 | -25.3 | -19.3 | -30.8 | -32.8 | ... |
| Māori Men All age | -16,000 | -12,220 | -19,510 | -20,790 | -171.9 | -131.3 | -209.6 | -223.3 | 3.7 |
| Non-Māori Men All age | -34,100 | -26,100 | -41,500 | -44,500 | -46.3 | -35.4 | -56.4 | -60.4 | ... |
| Māori Women All age | -1,900 | -1,390 | -2,450 | -2,470 | -18.0 | -13.2 | -23.2 | -23.4 | 3.4 |
| Non-Māori Women All age | -4,140 | -3,110 | -5,150 | -5,390 | -5.3 | -4.0 | -6.6 | -6.9 | ... |
| Māori Men 35-44 | -6,950 | -5,290 | -8,500 | -9,030 | -181.5 | -138.1 | -221.9 | -235.8 | 4.1 |
| Māori Men 45-54 | -6,080 | -4,590 | -7,550 | -7,900 | -179.9 | -135.8 | -223.4 | -233.7 | 3.7 |
| Māori Men 55-64 | -2,970 | -2,340 | -3,460 | -3,850 | -141.4 | -111.4 | -164.8 | -183.3 | 3.0 |
| Non-Māori Men 35-44 | -10,890 | -8,270 | -13,350 | -14,180 | -43.7 | -33.2 | -53.6 | -56.9 | ... |
| Non-Māori Men 45-54 | -12,870 | -9,650 | -16,090 | -16,760 | -48.4 | -36.3 | -60.5 | -63.0 | ... |
| Non-Māori Men 55-64 | -10,380 | -8,210 | -12,090 | -13,510 | -46.9 | -37.1 | -54.6 | -61.0 | ... |
| Māori Women 35-44 | 747 | 621 | 799 | 972 | 16.9 | 14.1 | 18.1 | 22.0 | 6.0 |
| Māori Women 45-54 | -1,265 | -952 | -1,573 | -1,648 | -33.1 | -24.9 | -41.2 | -43.1 | 5.9 |
| Māori Women 55-64 | -1,379 | -1,055 | -1,680 | -1,795 | -59.4 | -45.5 | -72.4 | -77.4 | 4.1 |
|  | Base case | Early recovery<br>in trade and<br>services | Extended border<br>control | Resurgence in<br>community<br>transmission | Base case | Early recovery<br>in trade and<br>services | Extended border<br>control | Resurgence in<br>community<br>transmission | Base case |
| Non-Māori Women 35-44 | 763 | 635 | 816 | 992 | 2.8 | 2.3 | 3.0 | 3.7 | ... |
| Non-Māori Women 45-54 | -1,563 | -1,167 | -1,964 | -2,037 | -5.6 | -4.2 | -7.0 | -7.3 | ... |
| Non-Māori Women 55-64 | -3,340 | -2,580 | -4,010 | -4,340 | -14.6 | -11.3 | -17.5 | -19.0 | ... |

**Notes:** All analyses in this table were implemented without uncertainty. Furthermore, please refer to the Additional scenarios and sensitivity analyses in the Methods section for changes in assumptions compared to the base-case analysis. The equity ratio for Māori/Non-Māori HALYs lost per capita was very similar for all scenarios.

**Supplementary Table 5.** Health loss (in HALYs) results for the base case (6% discount rate, Sensitivity analysis #E) for various unemployment scenarios.

| Groups | Change in HALYs |  |  |  | Change in HALYs per 1000 capita |  |  |  | Equity:<br>Māori/Non-Māori HALYs<br>per capita |
| --- | --- | --- | --- | --- | --- | --- | --- | --- | --- |
|  | Base case | Early recovery<br>in trade and<br>services | Extended border<br>control | Resurgence in<br>community<br>transmission | Base case | Early recovery<br>in trade and<br>services | Extended border<br>control | Resurgence in<br>community<br>transmission | Base case |
| All ethnicity Both sexes All age | -18,500 | -14,300 | -22,300 | -24,100 | -10.8 | -8.3 | -13.0 | -14.1 | ... |
| 5 years into future | -2,790 | -2,270 | -3,170 | -3,640 | -1.6 | -1.3 | -1.8 | -2.1 | ... |
| 10 years into future | -6,800 | -5,350 | -8,030 | -8,860 | -4.0 | -3.1 | -4.7 | -5.2 | ... |
| 20 years into future | -13,200 | -10,300 | -15,900 | -17,200 | -7.7 | -6.0 | -9.3 | -10.0 | ... |
| Māori Both sexes All age | -6,200 | -4,800 | -7,500 | -8,100 | -31.2 | -24.2 | -37.8 | -40.8 | 3.8 |
| Non-Māori Both sexes All age | -12,300 | -9,500 | -14,800 | -16,000 | -8.1 | -6.3 | -9.8 | -10.6 | ... |
| Māori Men All age | -5,500 | -4,250 | -6,630 | -7,150 | -59.1 | -45.6 | -71.2 | -76.8 | 4.0 |
| Non-Māori Men All age | -10,800 | -8,400 | -13,000 | -14,100 | -14.7 | -11.4 | -17.7 | -19.1 | ... |
| Māori Women All age | -720 | -540 | -910 | -940 | -6.8 | -5.1 | -8.6 | -8.9 | 3.6 |
| Non-Māori Women All age | -1,460 | -1,120 | -1,780 | -1,910 | -1.9 | -1.4 | -2.3 | -2.4 | ... |
| Māori Men 35-44 | -2,090 | -1,600 | -2,520 | -2,710 | -54.6 | -41.8 | -65.8 | -70.8 | 4.8 |
| Māori Men 45-54 | -2,160 | -1,640 | -2,650 | -2,800 | -63.9 | -48.5 | -78.4 | -82.8 | 4.3 |
| Māori Men 55-64 | -1,260 | -1,010 | -1,460 | -1,640 | -60.0 | -48.1 | -69.5 | -78.1 | 3.4 |
| Non-Māori Men 35-44 | -2,860 | -2,190 | -3,460 | -3,720 | -11.5 | -8.8 | -13.9 | -14.9 | ... |
| Non-Māori Men 45-54 | -3,990 | -3,020 | -4,940 | -5,200 | -15.0 | -11.4 | -18.6 | -19.6 | ... |
| Non-Māori Men 55-64 | -3,960 | -3,160 | -4,570 | -5,150 | -17.9 | -14.3 | -20.6 | -23.3 | ... |
| Māori Women 35-44 | 240 | 200 | 257 | 312 | 5.4 | 4.5 | 5.8 | 7.1 | 7.1 |
| Māori Women 45-54 | -409 | -311 | -504 | -534 | -10.7 | -8.1 | -13.2 | -14.0 | 6.6 |
|  | Base case | Early recovery<br>in trade and<br>services | Extended border<br>control | Resurgence in<br>community<br>transmission | Base case | Early recovery<br>in trade and<br>services | Extended border<br>control | Resurgence in<br>community<br>transmission | Base case |
| Māori Women 55-64 | -553 | -429 | -665 | -720 | -23.8 | -18.5 | -28.7 | -31.0 | 4.5 |
| Non-Māori Women 35-44 | 209 | 174 | 223 | 271 | 0.8 | 0.6 | 0.8 | 1.0 | ... |
| Non-Māori Women 45-54 | -455 | -342 | -565 | -592 | -1.6 | -1.2 | -2.0 | -2.1 | ... |
| Non-Māori Women 55-64 | -1,220 | -950 | -1,440 | -1,580 | -5.3 | -4.2 | -6.3 | -6.9 | ... |

*Notes: All analyses in this table were implemented without uncertainty. Furthermore, please refer to the Additional scenarios and sensitivity analyses in the Methods section for changes in assumptions compared to the base-case analysis. The equity ratio for Māori/Non-Māori HALYs lost per capita was very similar for all scenarios.*

